# Advice from a systems-biology model of the Corona epidemics

**DOI:** 10.1101/2020.03.29.20045039

**Authors:** Hans V. Westerhoff, Alexey N. Kolodkin

## Abstract

Using standard systems biology methodologies a 14-compartment dynamic model was developed for the Corona virus epidemic. The model predicts that : (i) it will be impossible to limit lockdown intensity such that sufficient herd immunity develops for this epidemic to die down, (ii) the death toll from the SARS-CoV-2 virus decreases very strongly with increasing intensity of the lockdown, but (iii) the duration of the epidemic increases at first with that intensity and then decreases again, such that (iv) it may be best to begin with selecting a lockdown intensity beyond the intensity that leads to the maximum duration, (v) an intermittent lock down strategy should also work and be more acceptable socially and economically, (vi) an initially intensive but adaptive lockdown strategy should be most efficient, both in terms of its low number casualties and shorter duration, (vii) such an adaptive lockdown strategy offers the advantage of being robust to unexpected imports of the virus, e.g. due to international travel, (viii) the eradication strategy may still be superior as it leads to even fewer deaths and a shorter period of economic lockdown maximum, but should have the adaptive strategy as backup in case of unexpected inflation imports (ix) earlier detection of infections is perhaps the most effective way in which the epidemic can be controlled more readily, whilst waiting for vaccines.

## Introduction

Different governments take different measures vis-à-vis the COVID-19 crisis, ranging from advice to reduce social activities, to a complete lock down of society and economy. Many governments do not seem to benefit maximally from the experiences of other countries. Almost invariably measures are taken too late. In this epidemic, times are too short for maximally informed, well-balanced deliberations leading to optimal and early conclusions. Policymakers, members of parliament, and voters, all require tools that enable them to anticipate better and to then fulfill their tasks. We here provide such a tool and we show how this tool leads to conclusions that could well prove crucial for managing the epidemic, as well as to a new adaptive control method.

## Methods

The construction of the diagram for the model of the COVID-9 epidemics used the open access CellDesigner software (v4.4; Systems Biology Institute, http://celldesigner.org/index.html). The network diagram was translated into a dynamic model using the open access software called COPASI (www.copasi.org) [2]. The resulting model was stored in the model/data repository FAIRDOMHub (http://doi.org/10.15490/fairdomhub.1.model.693.1) [3].

## Results

Using standard systems biology methodology we generated a dynamic model, which should apply to various countries after adjustment of the population size. For the unmanaged epidemic 3 % of the population is computed to die from COVID-19 infection within 3 months (Fig. 2). A ‘complete shutdown’ (modelled as a permanent reduction of infection probability by a factor of 10 through social distancing) should keep the % deceased down to 0.003 % (400 times lower than the natural death rate per year) [4]. A less complete lockdown leads to higher lethality in a highly nonlinear way (Fig. 3). Herd immunity is directly proportional to the percentage lethality at a proportionality constant of 27 (Fig. 3, and results not shown), so that in order to achieve the 50 % herd immunity that would quench this and a next epidemic, one would have to accept a death rate of 2 % in some 5 months, i.e. some three times the natural death rate: adjusting the lock down intensity so as to obtain sufficient herd immunity is unacceptable ethically.

**Fig. 1.**
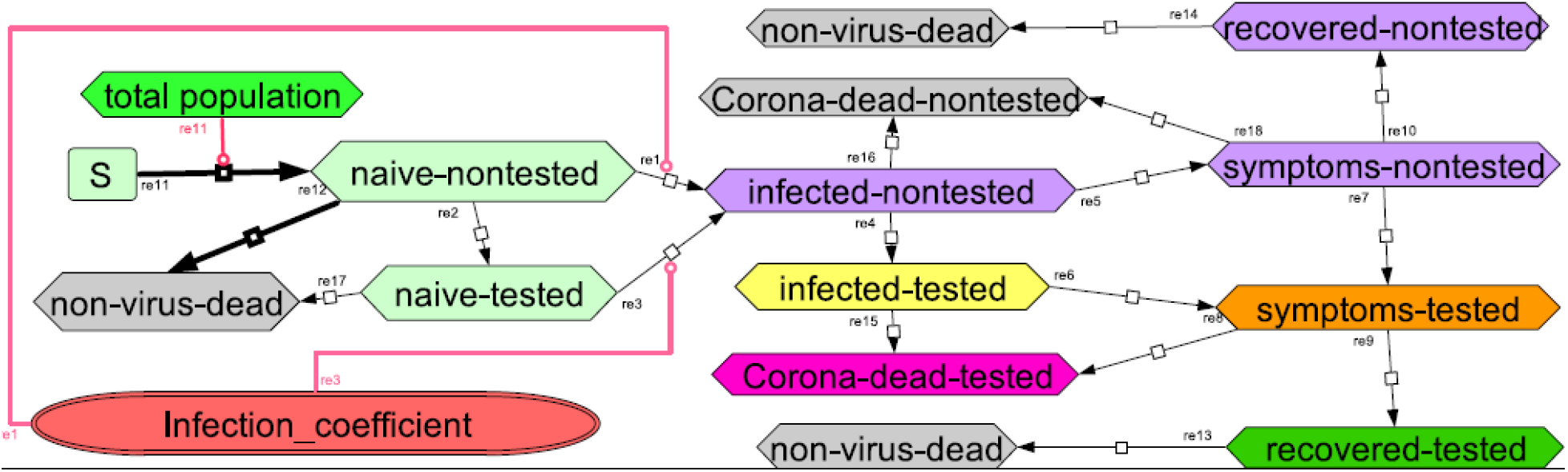
Systems Biology model of the Corona virus epidemics. Species are in boxes, reactions are indicated by arrows with reaction numbers written alongside. The infection coefficient is a linear function of the numbers of infected-nontested, infected-tested, symptoms-nontested and symptoms-tested, with relative weights 0.508, 0.25, 0.025 and 0.025 respectively divided by total population number and a social distancing factor. Model with all parameters values is at http://doi.org/10.15490/fairdomhub.1.model.693.1

**Fig 2.**
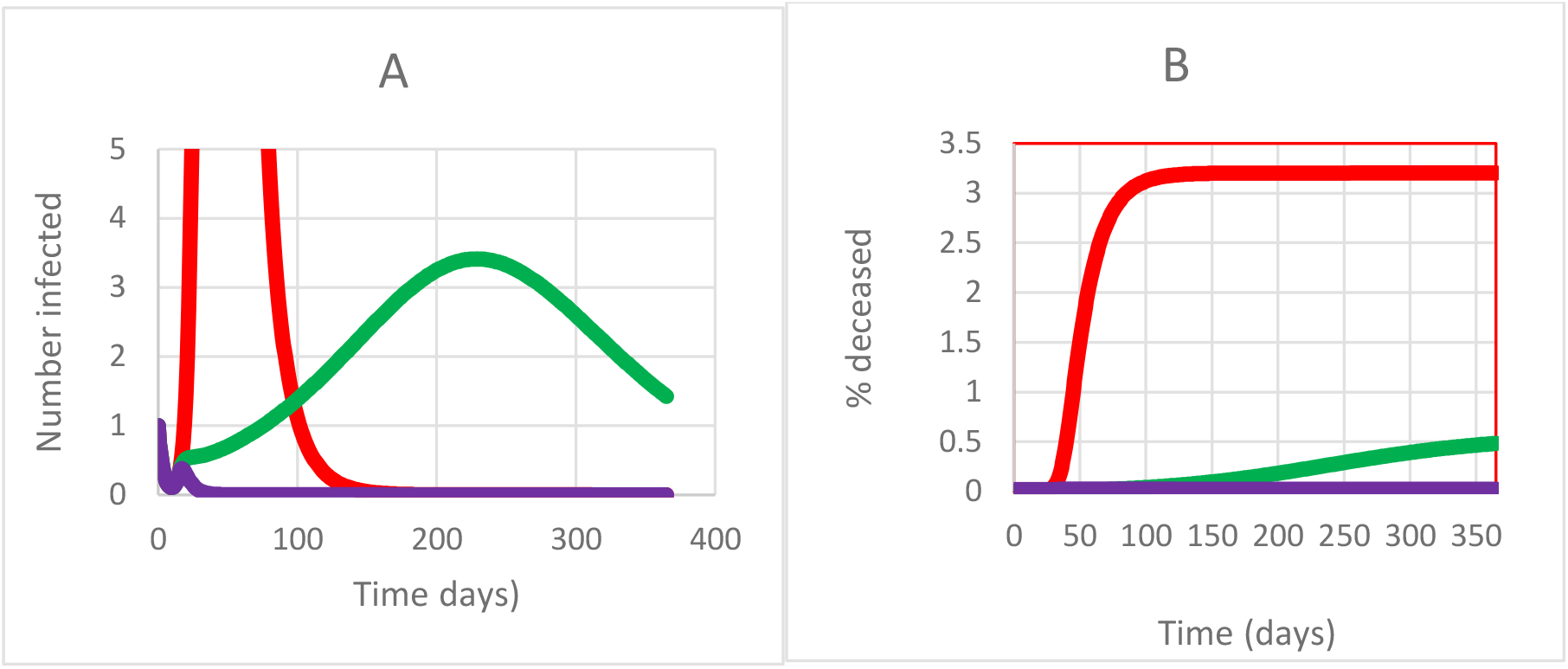
Predicted progression of epidemic in terms of (A) number of infected that test positively and (B) % deceased due to the virus; without government action (orange), with factor 2.2 social distancing (green) and with complete lockdown (blue).

**Fig. 3.**
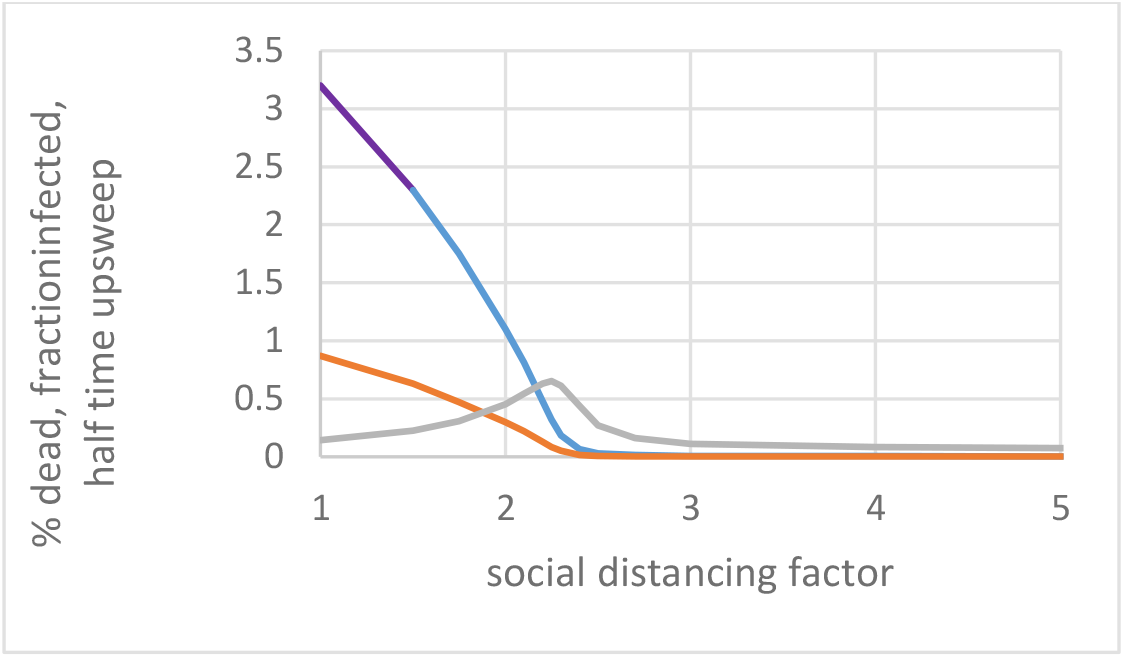
Effect of lockdown as function of lockdown intensity: % deceased (blue), fraction infected (orange), half rise time epidemic (in years; grey).

Acceptable strategies should perhaps focus on measures that keep the death toll around 0.03 %/year well below the natural death rate. Because the terminally ill occupy an intensive care bed for approximately a month with half of them surviving and assuming a peak lasting 3 months, this corresponds to approximately 2 such beds for every 10 000 inhabitants, close to the actual capacities in Northern Europe. A 2.5 fold permanent reduction in social interactions should reduce COVID-19 mortality from 3 % to this 0.03 % (Fig. 3). However, the duration of the epidemic (measured as the half time of its upsweep) varies appreciably with the social distance and around this 2.5 fold increase in social distance the duration of the epidemic appears to be the longest (Fig.3). This has the benefit of minimizing the challenge on the intensive care unit capacity, but might increase the economic damage.

A full lockdown should reduce the duration of the epidemic to 25 days, with a stronger but short effect on the economy (but see below for an extra strategy that will be needed to prevent re-emergence of the epidemic). On the other hand, a 3-fold decrease in social contact should already reduce the duration to 40 days (Fig. 3), with as additional benefit a much reduced lethality (0.008% compared with 0.03 at a 2.5 fold reduction). From this we conclude that a soft lockdown such as corresponding to a 2.5 fold reduction in social interaction could be the worst strategy to follow; a much stronger lock down is advisable. It should be noted that in none of these cases sufficient herd immunity will be reached to prevent a second epidemic.

A strong lockdown is hardship. Therefore we examined whether such a lockdown could be intermitted with periods with normal social contact, without endangering the success of the strategy. We found (Fig. 4A) that a 55%-on-45%-off schedule for the full lockdown will not suppress the epidemic. To suppress the SARS-CoV-2 virus two thirds of the time should be locked down, leaving one third for social interactions (Fig. 4B and supplementary material Fig. S1). This seems an attractive alternative to a permanent lock down provided a selection of economic activities that require live human-human interactions could be confined to shorter time periods without increasing contact intensity.

**Fig. 4.**
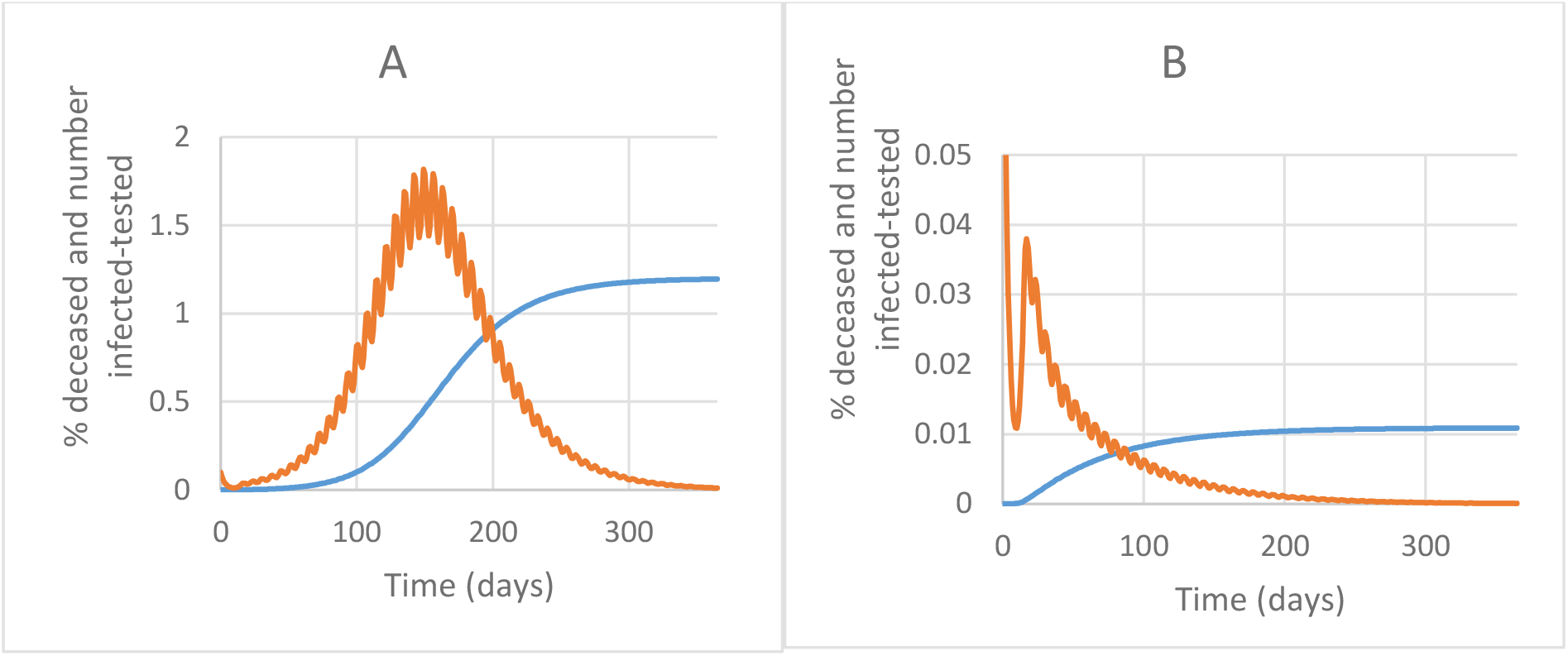
Intermittent lockdown scenarios; % death toll (blue) and number of infected-tested/10 (orange) as functions of time. (A) Intermittent lockdown 55% down/45% up and (B) Intermittent lockdown 70% down/30% up.

A more complex strategy should be one where the intensity of the shutdown is adapted to the severity of the epidemic. Choosing the fraction of the population that is newly tested as virus-positive as the controlling variable, this adaptive strategy should work better than a fixed lock down (Figure 5). Implemented at time 15 days after the first detection of an infected individual, this should lead to a lethality after one year of only 0.013 %, i.e. the one fourth the 0.33 % a continuous 2.25 fold lockdown would have led to (see Fig. 3). A disadvantage of this specific strategy is that the mortality increases linearly with time also after the first year. However, total mortality should not overtake the constant lockdown by 2.25 until after 10 years, long before which vaccines should be available. The adaptive lockdown could be optimized further in terms of parameters and with respect to any specific epidemic, culture and country.

**Fig 5.**
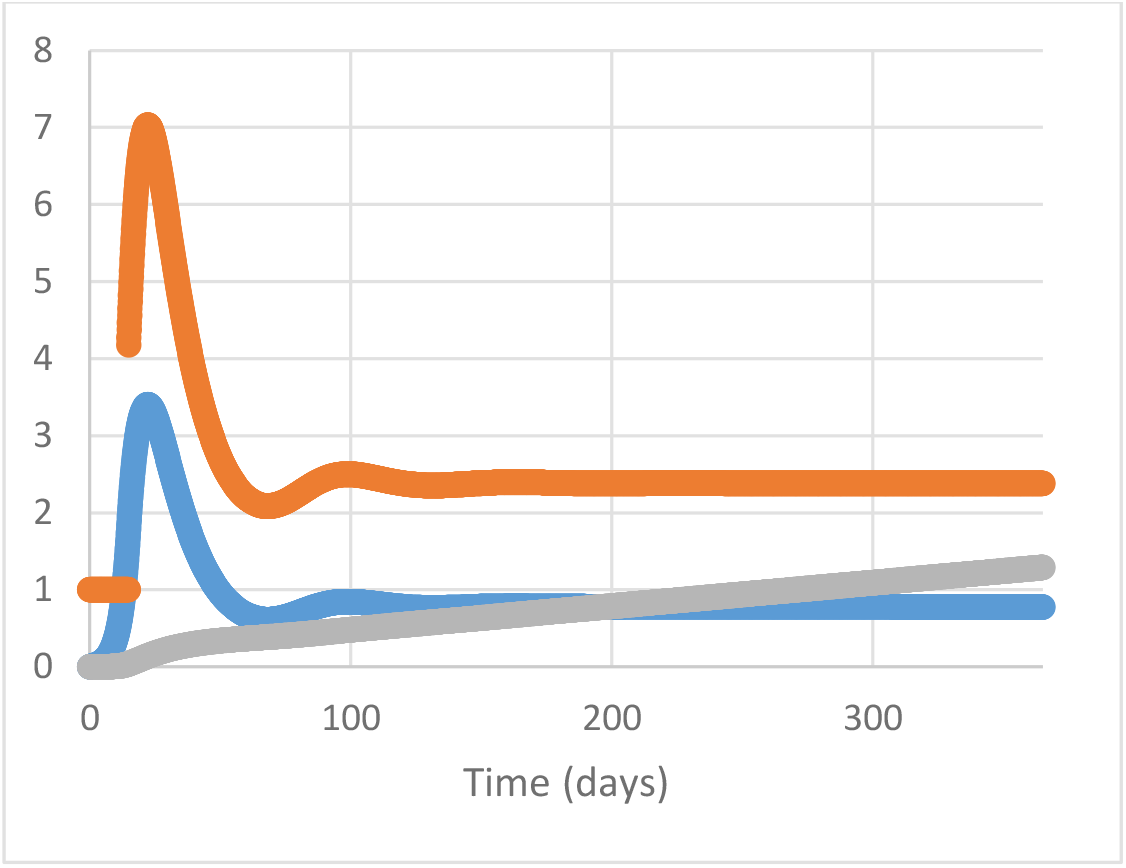
Adaptive lock down. Number of symptomatic tested (=controlling variable, /100, blue), Government enforced social distancing factor (orange) and % deceased (*100, grey line)

Figure 5 shows that for this adaptive strategy the effect on the number of individuals tested positively should be noticeable immediately after its onset: initially at least, the strategy has rather an intensive lockdown intensity, much stronger than what many governments are practicing. Most governments do the inverse: they exercise a soft lockdown first, increasing the lockdown intensity subsequently. The adaptive lockdown strategy proposed here begins with a strong lockdown to then relaxing it. The latter is the one that should work.

One of the important determinants of the success of a lockdown is how early in the epidemic it is enacted. This is even so for the adaptive lockdown strategy. Should the strategy be enacted 15 days later, then the number of dead at the end of the year becomes 20 times higher and the required initial lockdown a factor of 500, even though ultimately the lock down level should subside to the same factor of 2.2. So, governments should act earlier rather than later.

Faster testing of the symptomatic individuals should have little effect (results not shown), but a faster detection of symptoms in the individuals that have been infected should be highly effective: Doubling this rate constant reduces lethality after one year from 0.013 % to 0.0030 % and reduces the required social distancing peak to only 1.4. Quadrupling it reduces the distancing to a factor smaller than 1.1 and the death toll after a year to 0.00012 %. This same method will help detecting people that import the disease virus from abroad. Quarantine of the newly arrived persons should be effective, but should the entry of the infected people not be noticed, then the adaptive strategy should take care of it (results not shown), because the number of infected individuals is closely related to the control variable of the adaptive system.

An attractive alternative to the adaptive strategy is the full lockdown until virus extinction. Here the lock down strategy must be intensive and continued until there are no infected people left in the population (results not shown). However, this strategy is sensitive to import of new infections (Figure S2) and should only be robust if an adaptive strategy serves as back up.

The adaptive strategy does have the disadvantage that it should be maintained until a vaccine, or a much faster detection method of the infections, has arrived. Stopping the adaptive control at 180 days after onset, had the effect that the epidemic re-emerges with a half rise at 230 days, i.e. only one and a half month later (Fig. S3). The adaptive control method is incompatible with the extinction strategy however, as in the latter there is not control variable left. Yet the adaptive strategy can be used as back up.

## Discussion

Simulations using a quantitative model of the COVID-19 epidemic as a tool herewith showed that strategies aiming for herd immunity are unacceptable and that a much stronger lockdown is required. The argument that this strategy does not protect against imported virus infections, is irrelevant for this epidemic. Similar conclusions have also been achieved by more complex models [e.g. 4]. The advantage of the present model is that it is public domain and can be used by anyone with a personal computer. This paper further developed an adaptive strategy that should be superior over the lockdown strategy and deal with infections from abroad.

Our results suggest that the measures taken by many policy makers will be insufficient to quench the epidemic. Some Western policy makers engage in an adaptive lock down strategy but one of insufficient strength: our results suggest that their slowly increasing lock down strategy will not be effective. What is necessary is a strong lock down, which may then be softened as the number of infected individuals begins to decrease with time. Policy makers in other countries, such as in Taiwan and South Korea have been able to quench the epidemic in their country in this or harsher ways. The argument expressed by some European policy makers that such measures cannot be taken in democracies is thereby nullified. The Chinese measures appear to be closest to the full lock down strategy and have been most successful, but may also have been harsh on society. And certainly now that the remaining worry is import of new virus, China may be best off with quarantine of all visitors from abroad, with the strong adaptive strategy modeled here as a backup.

## Data Availability

The mathematical model used has been deposited in the corresponding database and is publicly accessible

http://doi.org/10.15490/fairdomhub.1.model.693.1

## Acknowledgements

We thank Marieke van Ham and Matteo Barberis for discussions.

## Supplementary material

**Figure S1.**
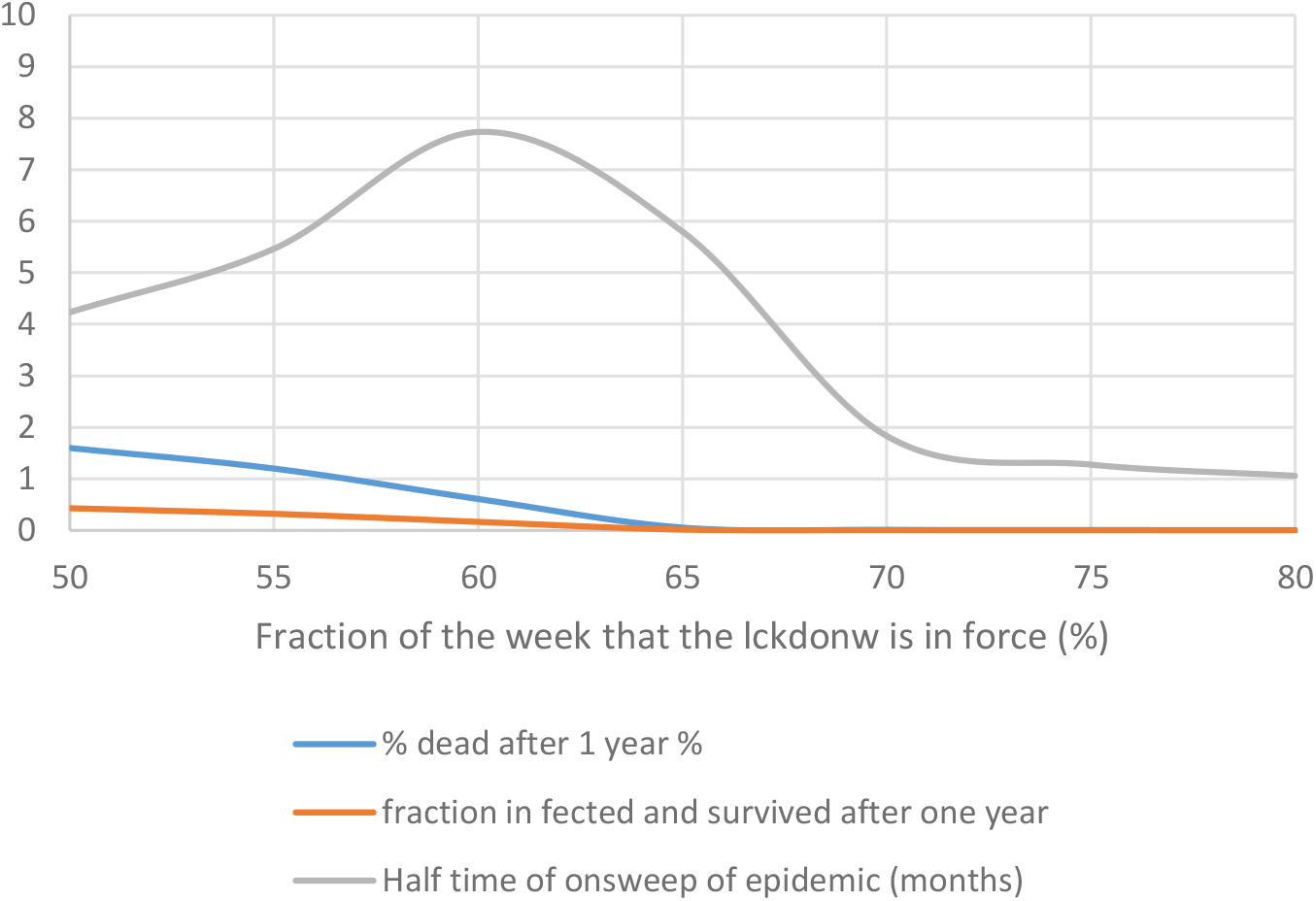
Effect of intermittent lockdown. Fraction dead, cured and epidemic rise time as function time fraction of intermittent lock down.

**Figure S2.**
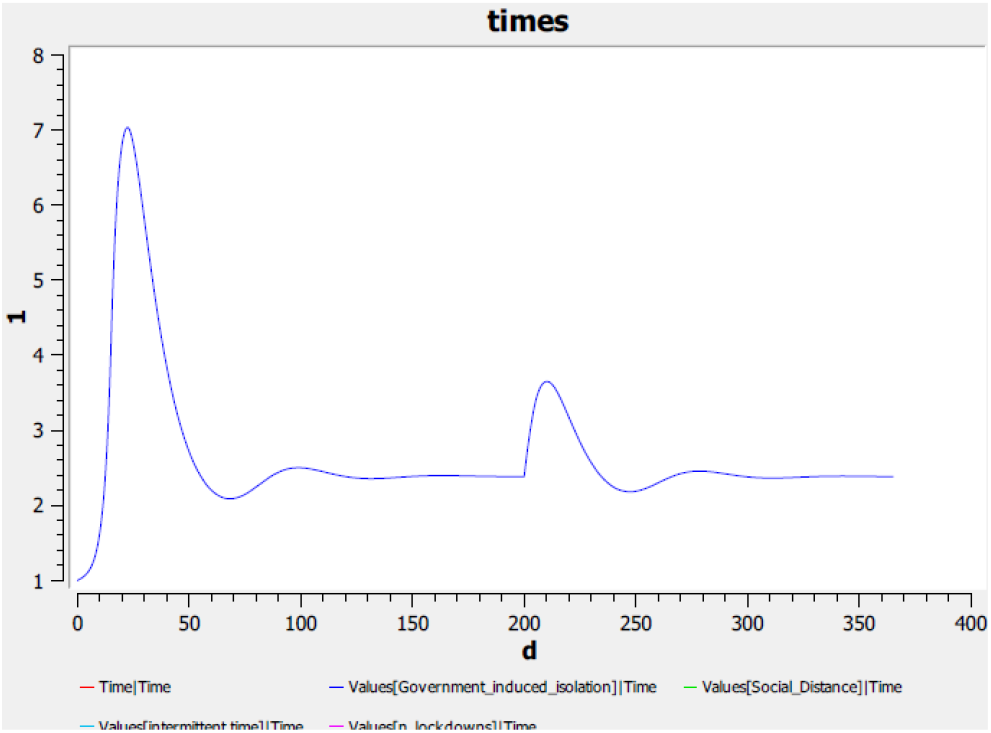
Foreign infection adaptive model. Government induced isolation factor is shown.

**Figure S3.**
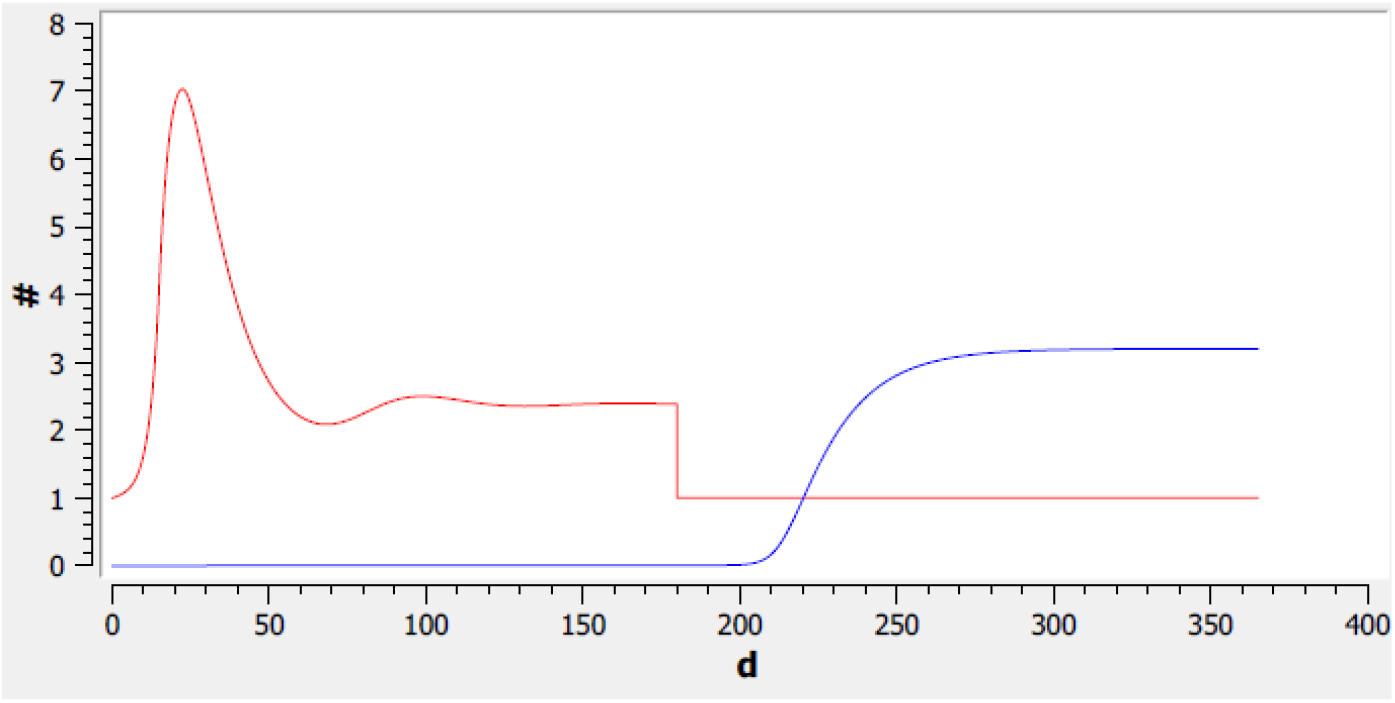
Epidemics when ending adaptive control at t=180. Blue: % deceased. Red: Adaptive control measure.

## References

1 Kitano, H., Funahashi, A., Matsuoka, Y. & Oda, K. Using process diagrams for the graphical representation of biological networks. Nature biotechnology 23, 961–966, doi:10.1038/nbt1111 (2005).

2 Hoops, S. et al. COPASI—a COmplex PAthway SImulator. Bioinformatics 22, 3074 (2006).

3 Westerhoff, H., & Kolodkin, A. (2020). Comprehensive model for SARS-CoV-2 virus spread in population, 2020 COVID-19 data. FAIRDOMHub. http://doi.org/10.15490/FAIRDOMHUB.1.MODEL.693.1

4 Ferguson, NM. et al. Impact of non-pharmaceutical interventions (NPIs) to reduce COVID-19 mortality and healthcare demand. Imperial College COVID-19 Response Team, March 16, 2020.

